# Impact of an ambient digital scribe on typing and note quality: the AutoscriberValidate study

**DOI:** 10.64898/2026.02.19.26346634

**Authors:** Martijn P. Bauer, Esther M. van Tol, Tarisha K.M. Constansia, Liza King, Marieke M. van Buchem

## Abstract

**Background:** Typing in the electronic health record (EHR) takes up healthcare providers’ time and cognitive space and constitutes a substantial administrative burden contributing to high burnout rates in healthcare. Ambient digital scribes may improve this problem.

**Objective:** To investigate the effect of the use of Autoscriber, an ambient digital scribe, on healthcare providers’ administrative workload and the quality of medical notes in the EHR.

**Methods:** A study period of 26 weeks was randomized into weeks when healthcare providers were allowed to use Autoscriber (intervention weeks) and weeks when they were not (control weeks) in a 2:1 ratio. Workload was assessed by comparing the number of characters typed in the medical note during control weeks with the number of modifications that were made to the summary produced by Autoscriber during intervention weeks. Quality of the medical note was measured by having a large language model (LLM) count the number of hallucinations, incorrect negations, context conflation errors, speculations, other inaccuracies, omissions, succinctness errors, organization errors and terminology errors per medical note.

**Results:** Between 1 November 2024 and 30 April 2025, 35 healthcare providers from 14 different specialties recorded 387 consultations in intervention weeks, and 142 in control weeks. The median number of characters typed per medical note was 1079 in control weeks and the median number of modifications necessary to produce the medical note was 351 in intervention weeks, compatible with a lower workload. All types of errors occurred significantly less frequently in notes made with the support of Autoscriber than in those without, except for speculations, where the difference did not reach statistical significance.

**Conclusions:** The use of Autoscriber resulted in a lower workload and a higher quality of the medical note.

## Introduction

The administrative burden on healthcare providers has increased in the past decades. One of the main contributors to this increasing administrative burden is the shift in documentation practices following the introduction of electronic health records (EHRs). (1-3) While EHRs offer advantages such as increased readability and accessibility of medical notes, they also contribute to an increased workload. Many healthcare providers feel that typing is too interfering with patient contact to do during the interaction with the patient, causing a shift of administration to after-hours. For many healthcare providers who do type during patient contacts, typing is taking up more cognitive space than writing used to do. This cognitively demanding task must be done simultaneously with other cognitively demanding tasks (retrieving information, building rapport with the patient, and producing a diagnostic and therapeutic plan). This may have a negative impact on the quality of the note and the other tasks. In addition, multiple other tasks, such as quality registration and entering billing codes, have come to contribute to the administrative burden. (4) EHRs can offer the advantage of reusable information, but in practice much of the information is entered in free-text fields making it difficult to reuse. Therefore, it is not surprising that the administrative burden was found to contribute to the high burnout rates in healthcare. (5)

Ambient digital scribes, software applications that capture relevant information from speech, have the potential to decrease the administrative burden of healthcare professionals. (6, 7) They may also make information more structured so that it can be reused for other purposes, such as ordering tests, communication with other healthcare professionals, quality registrations, etc.

One such ambient digital scribe is Autoscriber. We have previously conducted a study (8) using this tool to summarize recorded mock consultations. In this artificial setting, the support of Autoscriber reduced the time spent typing from 202 to 152 seconds, with comparable note quality. However, summaries were written by medical students without time constraint and without other tasks they had to perform.

The current study aimed to investigate the impact of Autoscriber in a real-world setting, focusing on workload and note quality.

## Methods

### Setting

The study was performed at the outpatient clinic and emergency department of the Leiden University Medical Center, an academic hospital of approximately 900 beds in the densely populated west of the Netherlands. Healthcare providers (physicians, physician assistants and nurse practitioners) across all medical specialties were invited to meetings where they received information on the study and could sign up to participate. Inclusion criteria for patients were age 16 or higher, the ability to provide informed consent and speaking Dutch. There were no inclusion criteria for healthcare providers, but we strove for an optimal balance between diversity of different specialties and at least two providers per specialty.

### Autoscriber

Autoscriber is a web-based software application that transcribes and summarizes medical conversations (currently with support for Dutch, English, German and Afrikaans). The initial stage of the pipeline relies on acoustic and transformer-based speech-to-text models to accurately convert spoken medical consultations into a speaker-diarized transcript. Following transcription, the application generates a structured clinical note or summary using a combination of large language models (LLMs) such as GPT-4o and Gemini 2.5. The system employs a modular prompt structure, tailored to specific medical domains, and performs several data quality validation and post-processing steps before the final summary is presented to the practitioner. This layered approach is designed to ensure that the output is medically accurate, coherent, and adheres to established clinical documentation stylistic guidelines. The application complies with the European Union General Data Protection Regulation (GDPR) and is certified according to the international ISO 27001 security standard as well as NEN 7510, the Dutch standard for information security in healthcare. Healthcare providers may use the application only after the patient has provided explicit verbal consent, which must be indicated before a recording may start. Upon ending the recording, the application displays the resulting transcript and summary within seconds. The healthcare provider has ultimate control over the final clinical note and is able to modify the summary at any point before transferring to and saving within the EHR. Modifications can be performed either directly within the Autoscriber application before export, or within the EHR system after initial transfer. Naturally, they may also choose to discard the generated summary entirely. To ensure data integrity, healthcare providers were instructed to make any necessary changes in the EHR and not in the application. This allows for assessing the quality of the non-redacted summary produced by Autoscriber, as well as analyzing the changes that the healthcare provider made to the Autoscriber summary.

### Procedures

The study followed a randomized design. For the 26-week study period, weeks were randomly assigned to intervention (Autoscriber) or control, in a 2:1 ratio. During intervention weeks, healthcare providers were allowed to use Autoscriber and copy the generated summary to the EHR. During control weeks, healthcare providers were not allowed to use Autoscriber to generate a summary. They were, however, asked to generate a transcript, so that this could serve as a reference when assessing the quality of the medical note. Healthcare providers were informed about whether a week would be an intervention week or a control week only the weekend before, to prevent selection bias from healthcare providers planning consultations in a specific week. All patients with a scheduled appointment received information about the study in advance. Healthcare providers were encouraged to include all patients in the study during the study period, but the decision to invite patients to participate in the study was ultimately left to the healthcare provider. In case of inclusion in the study, the healthcare provider obtained verbal informed consent before starting the consultation. During the first half of the study, patients who participated also received an anonymous questionnaire about how they perceived the contact with their healthcare provider.

### Study parameters

The primary study parameters were workload and the quality of the final medical note. Workload was assessed by the number of operations necessary to produce the medical note. For control weeks, this meant the number of characters that had to be typed. For intervention weeks, we assessed the Levenshtein distance (the number of single-character edits necessary to change one text to another) between the Autoscriber-generated summary and the final note in the EHR. Since the Levenshtein distance does not take operations like cut-and-paste into account, we also calculated the number of operations according to the diff-match-patch algorithm originally developed by the Google company (9). For practical purposes, only those parts of the medical note that can commonly be derived from a spoken conversation were considered, namely the medical history and the plan. Other parts of the note may be captured from a conversation, such as when a healthcare provider actively mentions their findings during a physical examination, but we felt that this would be too incidental to include in the current study. We also studied whether the workload was different for medical (defined as never working in an operating room, except for psychiatry), surgical (defined as sometimes working in an operating room) and psychiatric (defined as psychiatry) specialties. For the quality of the medical note, we considered the commonly used Physician Documentation Quality Instrument-9 and variants thereof. (10, 11) However, these metrics have been criticized for not weighing potential risks of errors. (12) In addition, the PDQI-9 is partly subjective, and inter-rater reliability may vary in our experience. We selected ten medical notes, had two raters rate the quality of the note using the PDQI-9 and calculated quadratic weighted Cohen’s kappa scores for seven attributes of the PDQI-9. Inter-rater reliability varied widely depending on the dimension (accuracy: 0.87, thoroughness: 0.61, usefulness: 0.35, organization: 0.09, comprehensibility: 0.70, succinctness: 0.22, internal consistency: 1.0). Therefore, we chose to use an LLM-based evaluation aimed to be as objective as achievable. The prompt instructs the LLM (GPT4o) to only consider parts of the transcript that may impact diagnosis or patient management decisions, and count the numbers of hallucinations (information presented as fact, but not present in the transcript), incorrect negations, context conflation errors (e.g., the patient mentions exposure to dust at home and to asbestos at work, and the LLM summarizes this as ‘exposure to dust and asbestos at work’), unwanted interpretations or speculations (statements that may be true, but are not fully justified based on the transcript), other inaccuracies (incorrect statements that cannot be categorized in the previous four categories), omissions, succinctness errors (unnecessary or redundant pieces of information, including duplications), organization errors (e.g., cigarette smoking mentioned in the social history when there is also a part about harmful substances elsewhere in the summary) and terminology errors (terms from the wrong register, e.g. medical jargon in the history of present illness, or layman’s terms in the physical examination) per transcript. The exact content of the prompt can be found in the Supplementary Materials. We found that calculating these LLM-generated counts ten times per summary and then averaging them resulted in consistent numbers and therefore used these counts.

Secondary study parameters were word error rate of the transcript, quality of the summary produced by Autoscriber, and patients’ perception of the contact with their healthcare provider, as assessed by the Dutch version of the Consultation and Relational Empathy (CARE) measure. (13)

To calculate the word error rate (WER), one researcher (MvB) listened to four recordings (a total of 57.1 minutes) and edited the Autoscriber transcripts, creating a set of gold standard transcripts. The WER was calculated by comparing the Autoscriber transcripts to the gold standard transcripts using Python version 3.8.5 and the ‘werpy’, ‘pandas’, and ‘spacy’ packages. Per recording, this led to a WER, the number of insertions, deletions, and substitutions.

### Sample size calculation

We aimed to include at least 20 consultations in the control arm and 20 in the intervention arm, which would result in a power of 89% to detect a 50-character difference (assuming a standard deviation of 50 characters), and a power of 89% to detect a 1-point difference in the numbers of each type of error in our quality evaluation (assuming a standard deviation of 1 points). Both power calculations were based on a desired two-sided confidence interval of 95%.

### Data analysis

Normally distributed data were summarized with means and standard deviations, non-normally distributed data with medians and interquartile ranges. Means of normally distributed data were compared using two-sided *t*-tests, non-normally distributed data were compared using two-sided Mann-Whitney *U*-tests. Statistical significance was declared at a *p*-value of 0.05 or lower. We used Python version 3.13.7 for the analysis, using the ‘pandas’, ‘numpy’, ‘scipy.stats’ and ‘sklearn.metrics’ packages.

### Ethical considerations

The study protocol was approved by the Scientific Committee of the department of Internal Medicine of the Leiden University Medical Center. This committee has been mandated by the Medical Ethics Board to assess protocols of studies that do not fall under the Medical Research Involving Human Subjects Act. The Scientific Committee judged the study not to fall under this act and as a consequence approval by the Medical Ethics Board was waived.

## Results

Thirty-five healthcare providers participated in the study. Between 1 November 2024 and 30 April 2025, these healthcare providers recorded 529 consultations, 387 in intervention weeks and 142 in control weeks. The specialties involved and the number of recordings per specialty are listed in Table 1. Despite the instruction not to generate a summary with Autoscriber during control weeks, healthcare providers sometimes generated a summary with Autoscriber, resulting in a total number of 456 Autoscriber-generated summaries. However, healthcare providers did not appear to use Autoscriber-generated summaries as a basis for their notes during control weeks. We included all Autoscriber-generated summaries in our analysis. The overwhelming majority (522, 99%) of recordings were made in the outpatient clinic or, in case of psychiatry, at the ward during an intake for admission. Seven recordings (4 by emergency medicine, 3 by general internal medicine) were made in the emergency department.

**Table 1:** Different participating specialties with the number of healthcare providers who participated and the number of recordings they made.

| Specialty | Healthcare providers | Recordings |
| --- | --- | --- |
| *Medical:* |  |  |
| Clinical genetics | 3 | 41 |
| Emergency medicine | 1 | 4 |
| Endocrinology | 3 | 35 |
| General internal medicine | 4 | 31 |
| Geriatrics | 1 | 2 |
| Nephrology | 2 | 74 |
| Neurology | 2 | 12 |
| Oncology | 1 | 17 |
| Rheumatology | 2 | 110 |
| Vascular medicine | 1 | 22 |
| Subtotal | 20 | 348 |
| *Surgical:* |  |  |
| Gynecology | 4 | 40 |
| Neurosurgery | 2 | 32 |
| Surgery | 3 | 29 |
| Subtotal | 9 | 101 |
| *Psychiatric:* |  |  |
| Psychiatry | 6 | 80 |
| Grand total | 35 | 529 |

### Workload

The results of the assessment of the workload are shown in Table 2. Healthcare providers used the Autoscriber-generated summary as a basis for their notes in intervention weeks but still redacted a lot of the text as shown by high Levensthein distances and number of operations according to the diff-patch-match algorithm for the comparison between Autoscriber-generated summaries and the final note in the EHR. However, the median number of diff-match-patch operations in intervention weeks was a third of the median number of characters typed in control weeks, suggesting a lower workload with Autoscriber than without.

**Table 2:**
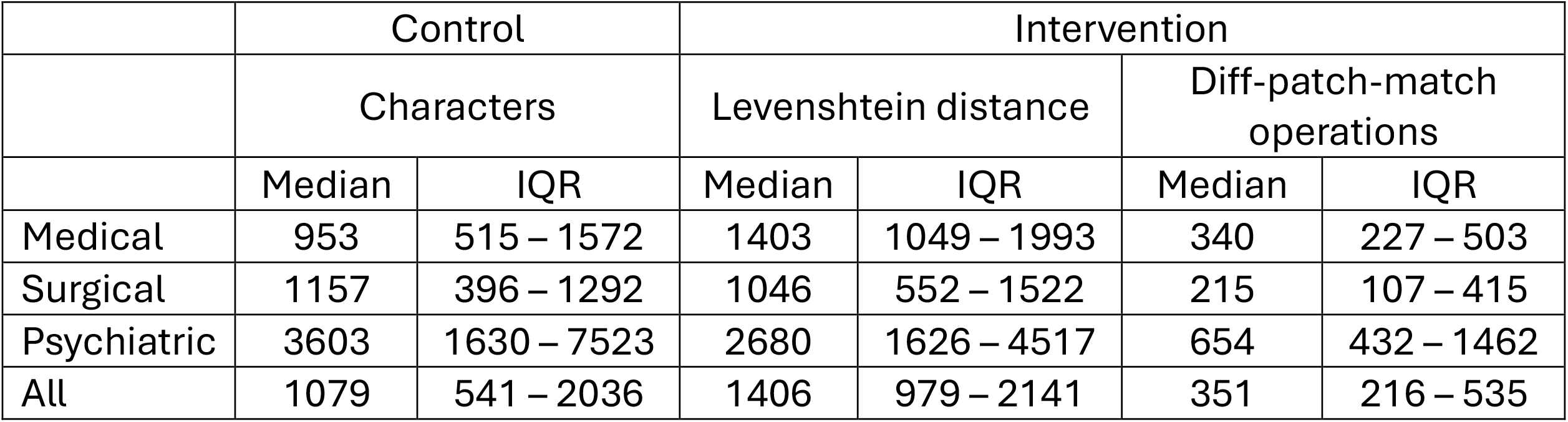
Workload as assessed by character counts in the medical notes made during control weeks and Levenshtein distance and operation count according to the diff-patch-match algorithm in the medical notes during intervention weeks. IQR = interquartile range.

|  | Control |  | Intervention |  |  |  |
| --- | --- | --- | --- | --- | --- | --- |
|  | Characters |  | Levenshtein distance |  | Diff-patch-match operations |  |
|  | Median | IQR | Median | IQR | Median | IQR |
| Medical | 953 | 515 – 1572 | 1403 | 1049 – 1993 | 340 | 227 – 503 |
| Surgical | 1157 | 396 – 1292 | 1046 | 552 – 1522 | 215 | 107 – 415 |
| Psychiatric | 3603 | 1630 – 7523 | 2680 | 1626 – 4517 | 654 | 432 – 1462 |
| All | 1079 | 541 – 2036 | 1406 | 979 – 2141 | 351 | 216 – 535 |

### Quality

The results of the assessment of the quality of the notes in control weeks and in intervention weeks are shown in Table 3, including a separate assessment of the quality of the Autoscriber-generated summaries. Notes created in intervention weeks were significantly better than those created in control weeks as assessed by the number of all types of errors, except for speculations, which were non-significantly less frequent in intervention weeks. Of note, hallucinations in control weeks probably do not reflect hallucinations in the sense of fully fabricated information presented as fact, but information that was simply not present in the transcript but known to the healthcare provider. Nonetheless, we have chosen to present hallucination counts in control weeks because we cannot distinguish between hallucinations and information known to the healthcare provider but not present in the transcript in notes created in intervention weeks and wanted to show the results in notes created in control weeks for comparison.

**Table 3:** Quality of summaries generated by Autoscriber and EHR notes in control and intervention weeks as assessed by counts of errors per summary or note. *P*-value for the comparison of notes in control weeks with those in intervention weeks. IQR = interquartile range.

| Errors per summary/ note | Autoscriber-generated summary |  | EHR note in control weeks |  | EHR note in intervention weeks |  | *p* |
| --- | --- | --- | --- | --- | --- | --- | --- |
|  | Median | IQR | Median | IQR | Median | IQR |  |
| Hallucinations/ information absent in the transcript | 0 | 0 – 1 | 2 | 1 – 27 | 1 | 0 – 2 | < 0.001 |
| Incorrect negations | 0 | 0 – 0 | 0 | 0 – 9 | 0 | 0 – 0 | < 0.001 |
| Context conflation errors | 1 | 0 – 1 | 1 | 0 – 20 | 0 | 0 – 1 | < 0.001 |
| Speculations | 1 | 0 – 2 | 1 | 0 – 2 | 1 | 0 – 1 | 0.06 |
| Other inaccuracies | 2 | 1 – 2 | 3 | 2 – 50 | 2 | 1 – 2 | < 0.001 |
| Omissions | 5 | 4 – 5 | 8 | 6 – 199 | 5 | 4 – 6 | < 0.001 |
| Succinctness errors | 3 | 2 – 4 | 4 | 2 – 63 | 2 | 1 – 3 | < 0.001 |
| Organization errors | 0 | 0 – 0 | 2 | 1 – 9 | 1 | 0 – 1 | < 0.001 |
| Terminology errors | 1 | 0 – 1 | 2 | 1 – 40 | 1 | 1 – 2 | < 0.001 |

### Secondary parameters

The overall word error rate (WER) was 0.05 (±0.01). See Table 4 for the scores per recording.

**Table 4:** Overview of the word error rate (WER), insertions, deletions, and substitutions, when comparing the gold standard transcriptions to the automatic transcriptions.

| Recording | Duration (minutes:seconds) | WER | Insertions | Deletions | Substitutions |
| --- | --- | --- | --- | --- | --- |
| 1 | 18:51 | 0.06 | 34 | 20 | 95 |
| 2 | 10:01 | 0.04 | 13 | 10 | 42 |
| 3 | 4:44 | 0.05 | 7 | 12 | 29 |
| 4 | 23:29 | 0.03 | 23 | 27 | 91 |

How patients viewed the contact with their healthcare provider was not significantly different during control weeks than during intervention weeks, as assessed by total CARE measure scores. The mean score was 43.73 ± 7.14 in intervention weeks and 44.47 ± 7.01 in the control weeks out of a maximum score of 50 (*p*-value 0.38).

## Discussion

In this study, we found that the use of Autoscriber reduced the amount of typing that healthcare providers must do for a consultation, while at the same time contributing to a better quality of the EHR note. In our previous study with mock consultations, we found a reduction in typing workload, but no improvement in note quality. (8) An explanation for this may be that the notes in that study were typed without time constraints or other sources of psychological stress by medical students who still wrote notes ‘by the book’, whereas the quality of real-world notes may be less optimal. (14) Previous studies have found mixed effects of digital scribes on time reduction. (15-18) Time gains were partly offset by the necessity to make edits to the notes generated by the software, as was also the case in our study. Previous studies have also provided some evidence that the quality of medical notes may be improved by digital scribes. (15) An interesting finding was the number of errors in human-generated notes in our study. The hallucinations we found in these notes were probably not hallucinations but rather information that was available to the healthcare provider but not present in the transcript. In contrast, the high amount of medically relevant information lacking from the note and other errors may reflect true lower quality of human-generated medical notes as compared to those made with the support of Autoscriber.

This study has several limitations. First, it was conducted in one academic hospital, and the findings may not be generalizable to other settings. Second, workload was measured by the proxy parameter of typing and operations like cut-and-paste. This parameter misses work added by correcting typos and work saved by pasting predefined text blocks. Ideally, workload would have been measured by a blinded observer clocking the amount of time spent administrating in the EHR, during the consultation and after the consultation, including during ‘pajama time’, when many healthcare providers finish their administrative tasks after hours. Unfortunately, we did not have the resources to do this. We considered the possibility of having healthcare providers measure their own administration time but felt this would be too burdensome for these healthcare providers and unreliable due to the lack of blinding. Third, we used a LLM to assess the quality of medical notes and Autoscriber-generated summaries, which may have not been representative of how humans would judge this. However, quality judgment by humans is not necessarily reproducible, showing subjectivity and inter-rater variation. (19) By using the most objective parameters we could think of, counts of specific error types, we hope to have produced a reliable quality metric. In addition, the LLM will not systematically favor notes written with the help of Autoscriber over those written without it, or vice versa. We also found that repeating assessment ten times and then averaging the scores made our quality metric highly reproducible. Fourth, our questionnaires to assess patient-perceived contact with the healthcare provider may not have been the optimal instrument to measure this. Since scores were very high both in control weeks and in intervention weeks, the instrument may have not been discriminative enough to capture subtle differences in eye contact and attention. A within-patient design, in which the same patient is assessed during both conditions, could reduce this bias in future studies. Also, it would have been interesting to assess healthcare providers’ perception of the contact with the patient. We felt systematically measuring this would have been too high a burden for participating healthcare providers. Anecdotally, healthcare providers reported making more eye contact with the patient, being able to focus better on the conversation, and being less fatigued at the end of an outpatient clinic. There is some support for these findings in previous studies. (15) Decreased fatigue may be explained by healthcare providers having one less task to perform during the consultation. In addition, correction of a text may be less cognitively demanding than generating a text from scratch.

In conclusion, our results indicate that support by an ambient medical scribe may reduce administrative workload for healthcare providers and improve the quality of the medical note. However, healthcare providers still typed a lot when they used Autoscriber. Personalization of how information is presented in the summary generated by Autoscriber, both stylistically and in terms of the organization of the note, might improve this. Future research should focus on what can be done to further reduce the amount of typing and on the effects of Autoscriber on healthcare provider-patient interaction, healthcare provider fatigue (and ideally burnout rate), and the quality of the diagnostic and therapeutic process. In addition, Autoscriber may capture information in a more structured way to be reused for test ordering, communication to other healthcare providers and other administrative tasks, thus contributing to a further reduction of administrative burden. This should also be a focus of further research.

## Data Availability

Access to our data is restricted because of their private nature. Readers can be provided an anonymized version of the dataset through contact with the corresponding author.

## End sections

### Funding

This study did not receive any funding.

### Authors’ contributions

Marieke van Buchem developed the idea for the study. Marieke van Buchem, Esther van Tol, Tarisha Constansia and Martijn Bauer wrote the study protocol. Esther van Tol and Tarisha Constansia conducted the study on an operational basis. Marieke van Buchem, Martijn Bauer and Esther van Tol performed the analyses. Martijn Bauer and Marieke van Buchem wrote the first draft of the manuscript. All authors read and improved the following versions of the manuscript.

## Abbreviations

CARE measure: Consultation and Relational Empathy measure.
EHR: electronic health record.
GDPR: European Union General Data Protection Regulation.
LLM: large language model.
WER: word error rate.

## Supplementary Materials

Prompt for calculating error counts per summary/ note:

“You are going to assess the quality of the summary of a transcript of a conversation between a healthcare provider and a patient. The text between the de triple backticks is the transcript of the conversation:

```{transcript}```

The text between the triple asterisks is a summary of the conversation.

***{summary}***

Only consider information in the transcript that should be captured in the summary: information that may impact diagnosis (previous diagnoses, symptoms with their corresponding features, exposure (to infection sources, medication, toxins, radiation, heat, pollutants, physical harm, life events and other psychological stressors), medication, allergies for and intolerance of medication and other substances, the occurrence of possibly heritable diseases in first- or second-degree biological family members, physical findings), other information that may impact patient management decisions (patient preferences, patient self-care, patient-reported quality of life, the patient’s support system), information on previous diagnoses and investigations that the patient gives to the healthcare provider, information that the healthcare provider gives to the patient on additional investigations, treatment options, prognosis, and the diagnostic and/or therapeutic plan that is decided upon. Count the number of hallucinations (defined as information presented as fact in the summary, but not present in the transcript). Count the number of incorrect negations (defined as facts are that are confirmed in the transcript, but negated in the summary). Count the number of context conflation errors (defined as correct pieces of information that have no relationship with each other in the transcript, but are connected to each other in the summary, for example, the patient mentions exposure to dust at home and to asbestos at work in the transcript, and this is summarised as ‘exposure to dust and asbestos at work’ in the summary). Count the number of unwanted interpretations in the summary (defined as information that may be correct, but cannot be stated as true without a doubt on the basis of the transcript, for example, the patient mentions smoking and frequent episodes of coughing in the transcript, but the summary mentions ‘frequent exacerbations of COPD due to smoking’). Count the number of omissions (defined as information present in the transcript, but absent in the summary). Count the number of succintness errors (defined as information in the summary that is redundant, duplicated or unnecessary considering the information that should be captured). Count the number of organisation errors (defined as correct information presented in the wrong part of the summary, for example, cigarette smoking is mentioned in the social history). Count the number of terminology errors (defined as parts of the summary that contain terms from the wrong register, for example, medical jargon is used in the history of present illness when the patient used layman’s terms, or layman’s terms are used in the physical examination). Present these numbers as follows: ‘Hallucinations: ‘, followed by the number of hallucinations, followed by ‘Incorrect negations: ‘, followed by the number of incorrect negations, followed by ‘Context conflation errors: ‘, followed by the number of context conflation errors, followed by ‘Speculations: ‘, followed by the number of speculations, followed by ‘Omissions: ‘, followed by the number of omissions, followed by ‘Succintness errors: ‘, followed by the number of succintness errors, followed by ‘Organisation errors: ‘, followed by the number of organisation errors, followed by ‘Terminology errors’, followed by the number of terminology errors, all on one line.”

